# The prevalence among heart failure patients and clinical profiles of patients with cor pulmonale at a tertiary center in Ethiopia

**DOI:** 10.1101/2024.08.16.24312095

**Authors:** Sewale Anagaw, Netsanet Fentahun, Habtamu Bayih, Yohannes Tekleab

## Abstract

**Background:** Heart failure is a complex clinical syndrome resulting from structural and functional impairment of ventricular filling or ejection of blood. Cor pulmonale is one type of this clinical syndrome. There are only a few published studies on cor pulmonale from Ethiopia. The objective of this study was to determine the prevalence among patients with heart failure and the clinical and laboratory profiles of patients with cor pulmonale who had follow up at one of the tertiary hospitals in Ethiopia

**Methods:** A hospital-based cross-sectional study was conducted at Felege Hiwot Referral Hospital from December 2018 to April 2019. A single population proportion formula was used to determine the number of heart failure patients that had to be included in the study to determine the prevalence of cor pulmonale. The medical records of cor pulmonale patients among the sample heart failure patients were then retrieved and data was extracted using a structured checklist. Data was entered into, cleaned, and analyzed using IBM.SPSS version 23.0. Descriptive statistics were used to report the findings.

**Results:** Eight percent (35) of patients with heart failure had cor pulmonale. Fifty-four point three percent (19) of the patients with cor pulmonale were males and 45.7 %(16) were females. The median age of patients with cor pulmonale was 55 years. The commonest clinical features were cough and dyspnea (91.4 % and 97.1 % respectively). All patients had right ventricular dilation on echocardiography. Pulmonary Complications post-treatment for tuberculosis were the leading causes followed by interstitial lung disease. There was no identified respiratory disease in 40% of patients with cor pulmonale

**Conclusion:** Cor pulmonale accounted for less than 10 % of heart failure cases. Complications post-pulmonary tuberculosis were found to be the leading respiratory conditions underlying the cor pulmonale. Programs on prevention, early detection, and treatment of pulmonary tuberculosis must be strengthened.

## Introduction

Heart failure (HF) is a complex clinical syndrome resulting from structural and functional impairment of ventricular filling or ejection of blood. It is a burgeoning problem worldwide, affecting more than 20 million people (1). The overall prevalence of HF in the adult population in developed countries is 2%. Population-based studies on the incidence and prevalence of HF in developing countries are evolving. It is estimated that cardiovascular (CV) diseases accounted for 7%–10% of all medical admissions to African hospitals and HF constitutes 3%–7% of these admissions (2). More than two-thirds of all cases of HF can be attributed to four underlying conditions: ischemic heart disease, chronic obstructive pulmonary disease ending up in core pulmonale, hypertensive heart disease, and rheumatic heart disease (3, 4, 5).

The term cor pulmonale is still popular in the medical literature, but no consensual definition exists. Some define cor pulmonale as the alteration of right ventricular structure or function that is due to pulmonary hypertension (PH) caused by diseases affecting the lung or its vasculature (excluding right-sided heart disease from primary disease of the left side of the heart or congenital heart disease) (6). Others define it as PH resulting from diseases affecting the structure and/or the function of the lungs; PH results in right ventricular enlargement and may lead with time to right heart failure (RHF) (7). Reliable estimates of the prevalence of cor pulmonale are difficult to obtain because of this variability in definition (8). In many countries in Africa, the reported prevalence amongst cardiac patients ranges between none to 15% (9). To the investigators’ knowledge, there were only a few published studies on cor pulmonale in Ethiopia till the time of the study, and there was no single study that specifically assessed the proportion of cor pulmonale among all heart failure cases. This study determined the proportion of patients with heart failure who had cor pulmonale, and it also assessed the clinical and laboratory profiles of patients with cor pulmonale.

## Materials and methods

### Study Area and Period

A hospital-based cross-sectional study was conducted at Felegehiwot Referral Hospital (FHRH), a tertiary hospital located in Bahirdar town, Ethiopia. Bahirdar is the capital city of Amhara National Regional State and it is located at an altitude of 1,820 meters above sea level, 578 kilometers northwest of Addis Ababa. The hospital started giving service in 1955 and it was initially built to serve around 25,000 people. At the time of the study, it had 9 wards and 435 beds for inpatient management and outpatient clinics, serving an estimated 5-7 million people (10). The Department of Internal Medicine at FHRH has inpatient and outpatient units. The outpatient unit included follow-up clinics for chronic medical conditions like diabetes mellitus and heart failure. The chronic follow-up clinic was staffed with internists, internal medicine residents, and general practitioners. The study was conducted at the follow-up clinic between December 2018 and April 2019.

### Sampling method and data analysis

The Study population were all patients with heart failure who had a follow-up visit to FHRH chronic follow-up clinic in the study period mentioned. Patients with objective evidence of heart failure were included, and those who were labeled as cases of heart failure in their medical records but didn’t have radiographic and echocardiographic evaluation or evidence were excluded. Single population proportion formula was used to calculate the sample size necessary to determine the prevalence of cor pulmonale patients among heart failure patients. Samples were then selected using convenience sampling technique. The final sample size required was calculated to be 433 after correction for finite population and possible missing data. Medical records of those selected 433 heart failure patients were retrieved using their medical registration number and then medical records of cor pulmonale patients among those sample heart failure patients were placed separately for data extraction on clinical and imaging profiles. A structured checklist was used to extract data on clinical and laboratory profiles of cor pulmonale patients. The checklist was developed based on previous studies and modified based on the objective of the current study. It had 3 parts: part one was about socio-demographic and clinical characteristics, part two about imaging findings, and part three about underlying respiratory diseases. Data was entered into, cleaned, and analyzed using IBM.SPSS.STATISTICS version23. Means were used for scale variables and percentages were used for nominal variables.

### Ethical considerations

Ethical approval from the management board of Felegehiwot referral hospital was sought and a written approval paper was taken from the board before data collection was started. Medical records (hard copies) were accessed in the study period after the ethical approval letter was submitted to the registration and recording unit of the outpatient department of the hospital where medical records kept. Each participant’s information was collected using an anonymous checklist. The authors didn’t have any access to information that could identify individual participants during or after data collection

### Operational definitions

**Heart failure:** a diagnosis of heart failure was considered if there were documented symptoms (dyspnea, orthopnea, paroxysmal nocturnal dyspnea, leg and abdominal swelling, or fatigue), signs (leg edema, raised jugular venous pressure, rales, accentuated P2, or shifted point of maximal impulse), radiologic evidence (pulmonary edema, cardiomegaly) and echocardiographic findings suggestive of ventricular dysfunction (1, 3).

**Cor pulmonale:** diagnosis of core pulmonale was made if there was a documented echocardiography that showed pulmonary hypertension (estimated systolic pulmonary arterial pressure ≥ 40 mmHg, or at least moderate if no objective record of pulmonary arterial pressure) with concomitant alteration of right ventricular structure and/or function, and no evidence of primary left-sided heart disease and congenital heart disease (6).

**Electrocardiographic (ECG) evidence for core pulmonale**: documented ECG findings taken as evidence for cor pulmonale were right axis deviation, p pulmonale in limb lead II or III, R/S amplitude greater than1 in lead V1, and poor R wave progression (6,7)

**Plain chest radiography (CXR) evidence for core pulmonale**: enlargement of the central pulmonary arteries, cardiomegaly, and/or signs of underlying lung disease were taken as evidence for cor pulmonale (7)

**Echocardiographic evidence for core pulmonale:** findings considered diagnostic for cor pulonale on two-dimensional transthoracic echocardiography were right ventricular hypertrophy, right ventricular dilatation, reduced right ventricular function, and tricuspid regurgitation (11)

**Polycythemia:** Hemoglobin levels of 16.5g/dl for females and 18.5g/dl for males were used as cut-off for diagnosis of polycythemia.

**Diagnosis of underlying respiratory conditions**: Diagnosis of obstructive airway disease was considered if the documentation on the medical record of the patients showed a history of asthma/chronic obstructive pulmonary disease (COPD) on treatment, wheezes on examination, and hyper-inflated chest x-rays with or without spirometric evidence. Interstitial lung disease (ILD) was taken as the underlying respiratory disease if there was documentation of dyspnea with interstitial opacities and/or honeycombing on chest imaging (computed tomography or CXR). If the patient had a documented history of treatment for pulmonary tuberculosis at any point in time throughout his/her life and radiographic abnormalities on chest imaging (fibrosis, lung volume loss, bronchiectasis, or any combination of these), the label for the underlying structural lung disease would be pulmonary complication post-treatment for tuberculosis. The underlying respiratory condition was labeled as unidentified when chest X-ray showed no abnormality other than cardiomegaly and/or enlargement of central pulmonary arteries, chest CT is either normal or not done and spirometry is again either normal or not done

## Results

Thirty-five (35) of the 433 patients with heart failure (8.08 %) had cor pulmonale

### Socio-demographic and clinical characteristics of cor pulmonale patients

Nineteen (54.3 %) of patients with cor pulmonale were males and 45.7 % were females. The median age was 55 years (range 27-84 years), and 75% of them were below 66 years of age. Ninety-four-point three percent (33) came from rural areas. The majority of patients (80%) didn’t have documentation on their occupation. All of the patients who had documentation on their occupation were farmers. Ninety-one point four percent complained of cough, and 97.1 % had dyspnea. Leg edema was seen in 71.4 % and hepatomegaly in 37.1%. Only 28.6 % of patients had ascites, and wheezing was heard on auscultation in 14.3 % of patients. The majority (82.9% for cyanosis and 94.3% for clubbing) of them did not have a record of the presence or absence of cyanosis and clubbing. Cyanosis was seen in 33.3% and clubbing in 50 % of cases who had documentation of cyanosis and clubbing. Forty-two-point nine percent (15) had accentuated P2.

**Table 1:**
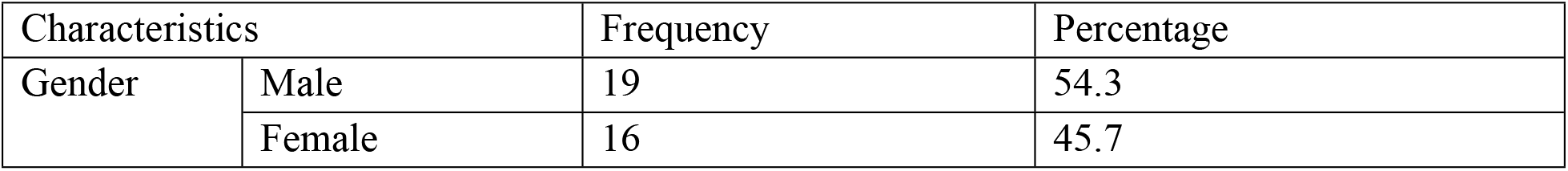
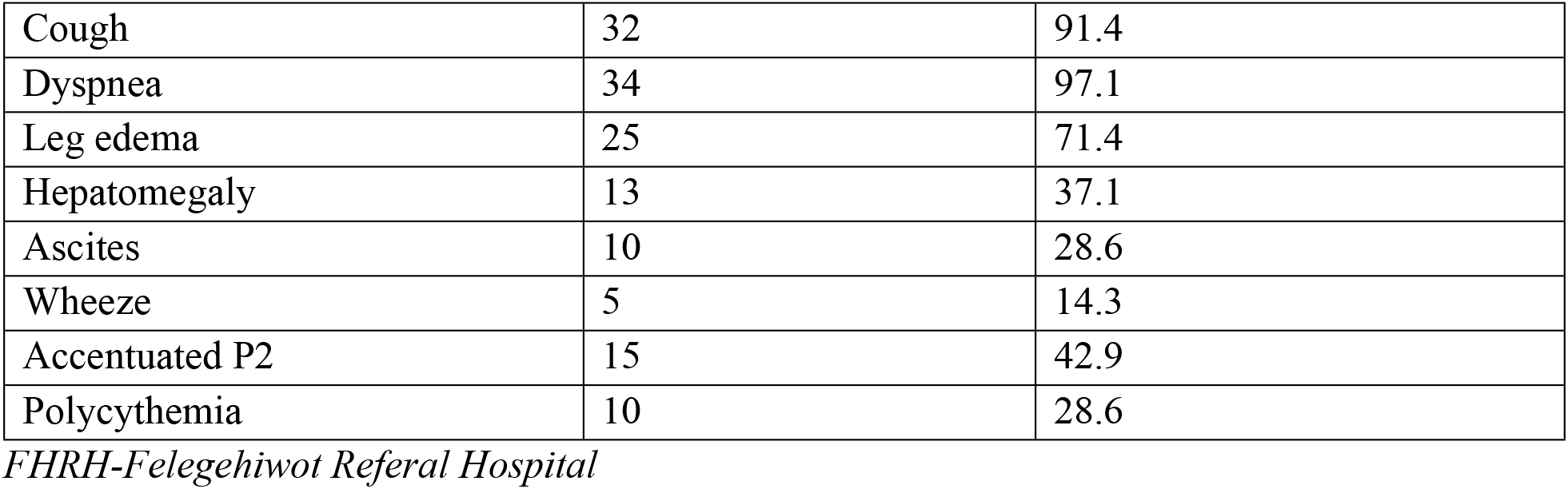
Summary of socio-demographic and clinical characteristics of cor pulmonale patients (n=35) at FHRH, Bahirdar Ethiopia, 2019

| Characteristics |  | Frequency | Percentage |
| --- | --- | --- | --- |
| Gender | Male | 19 | 54.3 |
|  | Female | 16 | 45.7 |

|  |  |  |
| --- | --- | --- |
| Cough | 32 | 91.4 |
| Dyspnea | 34 | 97.1 |
| Leg edema | 25 | 71.4 |
| Hepatomegaly | 13 | 37.1 |
| Ascites | 10 | 28.6 |
| Wheeze | 5 | 14.3 |
| Accentuated P2 | 15 | 42.9 |
| Polycythemia | 10 | 28.6 |

*FHRH-Felegehiwot Referral Hospital*

### Laboratory and imaging findings

Twenty-eight-point six percent (10) had polycythemia. The mean hemoglobin was 19.3 g/dl (SD ±1.57) (range 17-22 g/dl). Mean hemoglobin was higher among males compared with females (20.2 g/dl versus 18.2g/dl). ECG evidences were seen in 57.1 % of cases. Seventeen-point one percent (6) did not have evidence of cor pulmonale on ECG and there was no documented ECG in 25.7 % of cases. Among those who had ECG abnormalities, 20% had all the features (right axis deviation, P pulmonale in leads II/III, R/S greater than 1 in lead V1, and poor R wave progression) and 65 % had more than one feature of cor pulmonale. Fifteen percent of those with ECG features of cor pulmonale had only a single evidence: 5% only right axis deviation, 5% only P pulmonale and the other 5% had only R/S amplitude greater than 1 in lead V1.

Chest x-ray was normal in 5(14.3 %) patients. Two patients did not have chest X-rays. Twenty-eight (80%) patients had abnormal chest x-ray with findings suggestive of heart failure and /or underlying respiratory disease. Among those who had abnormal chest x-rays, 12 (42.9%) had only evidence of heart failure(cardiomegaly with or without enlargement of central pulmonary arteries), 12(42.9%) had evidence for both heart failure and underlying respiratory disease and the remaining 4(14.2%) patients had only evidence of underlying lung disease.

Six patients (17.1%) underwent chest computed tomography (CT) imaging, and it was abnormal in all of those patients. One of them was a known systemic sclerosis patient. CT showed basal and subpleural honeycombing with mild traction bronchiectasis. The other patient had bilateral asymmetric predominantly upper lung zone honeycombing with ground glass opacity and traction bronchiectasis. Bilateral fibrotic changes with bilateral multifocal pleural thickening and honeycombing, right upper lung fibrosis and emphysematous changes with bullae, bilateral end-stage fibrosis with honeycombing, and basal bronchiectasis were CT findings in the remaining four patients. Only a single patient had spirometry and the finding was that of obstructive pattern. All patients had right ventricular (RV) dilation on two-dimensional transthoracic echocardiography. Thirty-four (97.1%) patients had tricuspid regurgitation and 3 (8.57%) patients had right ventricular wall hypertrophy in addition to RV dilation. RV systolic function was assessed in 21 (60 %) of patients and it showed reduced function in 8. Estimated systolic pulmonary arterial pressure was reported as severe in one patient and moderate in four patients. In the remaining 30 patients, it was reported quantitatively. The mean estimated pulmonary arterial pressure was 71.5mmHg (SD±19.1) (Range 45-120 mmHg).

**Table 2:** Abnormal lung parenchymal findings reported on CXR among patients with cor pulmonale (n=16) at FHRH, Bahirdar Ethiopia, 2019

| Finding | Frequency |
| --- | --- |
| Fibrotic changes | 4 |
| Bronchiectasis | 1 |
| Interstitial opacities | 4 |
| Fibrosis with lung volume loss | 1 |
| Fibrosis, lung volume loss, and bronchiectasis | 2 |
| Cavitary lesions | 2 |
| Hyperinflation | 1 |
| Kyphoscoliosis | 1 |

*CXR- chest X-ray; FHRH- Felegehiwot Referral Hospital*

### Underlying respiratory diseases

Forty percent (14) of patients did not have any identified parenchymal lung disease as the underlying etiology. Pulmonary complications post -post-tuberculosis, ILD, obstructive airway disease, kyphoscoliosis, bronchiectasis, and connective tissue disease-related respiratory disease were the identified disease entities in order of decreasing entities.

**Table 3:** List of identified underlying respiratory diseases among patients with cor pulmonale at FHRH, Bahirdar, Ethiopia, 2019

| Disease | Frequency | Percentage |
| --- | --- | --- |
| obstructive airway disease | 4 | 11.4 |
| ILD | 5 | 14.3 |
| Complications post pulmonary tuberculosis | 8 | 22.9 |
| Bronchiectasis | 1 | 2.9 |
| Kyphoscoliosis | 2 | 5.7 |
| Connective tissue disease | 1 | 2.9 |
| Not identified | 14 | 40.0 |
| Total | 35 | 100 |

*ILD- Interstitial lung disease*

## Discussion

After the age of 50 years, cor pulmonale is the third most common cardiac disorder, after coronary and hypertensive heart disease in the USA based on a study done at the University of Pennsylvania (8). Cor pulmonale accounted for 7.7% in Kenya,1.6-4.4% in Nigeria,2.4% in Ghana, and 2.1-2.7% in Togo, of heart failure cases based on a systematic review published in the International Journal of Cardiology (12). A hospital-based study in South Africa showed that cor pulmonale is responsible for 11.8% of heart failure cases (13). In Ethiopia, a study done at Addis Ababa University reported that 1.6% of cardiac patients had cor pulmonale (14). The proportion found in this study (8.08 %) is within the range of 0-15% reported for most African countries. Unlike this study, the definition and diagnostic parameters used for the diagnosis of cor pulmonale were not clearly stated in the aforementioned reports. Therefore, comparing the prevalence in this study with the above reports could be difficult. Besides this, the study done at Addis Ababa University used only CXR and ECG for the diagnosis of cor pulmonale.

There are limited studies both worldwide and nationwide on the clinical characteristics of patients with cor pulmonale. A median age of 55 years found in this study was close to the median age of 51 reported from another study in Ethiopia (9). The prevalence of secondary polycythemia was reported to be 80 % in the above study (compared with only 28.6% in our study), but the mean hemoglobin levels were comparable (18.3 g/dl in the previous study versus 19.3 g/dl in the current study). A study done at a tertiary center in India showed that cough and dyspnea were the two commonest manifestations, and these manifestations were found to be the commonest ones in our study too (15). The prevalence of clubbing was higher (70%) in the Indian study compared with the prevalence in this study(50%), but the sample size difference (60 in the Indian study versus 35 in the current study) and lack of a complete record on clubbing in our study might have accounted for the lower prevalence in this study.

The top three causes of cor pulmonale reported in Ethiopia are COPD, bronchial asthma, and pulmonary tuberculosis-related complications (9). Similar causes have been identified in our study, although COPD and bronchial asthma were grouped into one in the medical records. Interstitial lung disease (ILD) was more prevalent in our study compared with a previous similar study in Ethiopia (14.3 % Vs 11%). A Kenyan case-control study among women with isolated right-sided heart failure (IRHF) (synonymous with our definition of cor pulmonale) showed that lower kitchen ventilation and occupational dust exposure were associated with IRHF (16). Most of the patients in our study came from rural areas and thus factors found to be associated with IRHF in the Kenyan study might have contributed to the cor pulmonale prevalence in our study but prospective studies are required to make such conclusions.

The major limitation of this study is that patients were not directly examined by the data collectors; rather, secondary data was taken from the medical records of patients. This makes the study results dependent on the completeness of prior patient evaluation and documentation. The other limitation is that the sample size for cor pulmonale was small. We believe that some of the underlying respiratory diseases might be underrepresented and others overrepresented in our study. For instance, in resource-limited settings like ours, there is a higher tendency to try patients with chronic cough with anti-tuberculosis drugs even if there is no microbiologic evidence and this might have contributed to our study result. This study has provided detailed data on the clinical presentation and imaging findings of patients with cor pulmonlae which can be used as an entry point for future studies. Prospective studies with clinical evaluations by data collectors well trained for this purpose and investigations directed at underlying causes of cor pulmonale are required to better define the characteristics and causes of cor pulmonale.

## Conclusion

Cor pulmonale accounted for 8.08 % of heart failure patients in this study. Most of the patients came from rural areas. Cough and dyspnea were the most common clinical manifestations and right ventricular dilation was the universal finding on echocardiography. The investigations directed at underlying causes of pulmonary hypertension were limited. Only limited patients had undergone chest CT and spirometry. Tuberculosis-related complications were the leading respiratory diseases followed by interstitial lung disease and obstructive airway disease. Patients with cor pulmonale need to be worked up well to identify the underlying respiratory diseases that could be manageable. National programs on prevention, early detection, and treatment of pulmonary tuberculosis must be strengthened.

## Data Availability

data will be provided by the corresponding author on request

## Acknowledgment

We would like to thank Bahirdar University College of Medicine and Health Sciences for the financial support. The study was fully funded by this college.

